# Substance P, mast cells and basophils are involved in acute chest syndrome in sickle cell disease

**DOI:** 10.64898/2026.03.02.26347450

**Authors:** Slimane Allali, Rachel Rignault-Bricard, Christine Ibrahim, Geoffrey Cheminet, Sarah Mattioni, Jacques Callebert, Aline Santin, Romain Fauchery, Marie Bouillié, Jean-Benoit Arlet, Valentine Brousse, Joséphine Brice, Mariane de Montalembert, Claire Heilbronner, Jean-Marie Launay, Sophie Georgin-Lavialle, Olivier Hermine, Thiago Trovati Maciel

**Affiliations:** Department of General Pediatrics and Pediatric Infectious Diseases, Reference Center for Sickle Cell Disease, Necker-Enfants Malades Hospital, Assistance Publique – Hôpitaux de Paris (AP-HP), Université Paris Cité, Paris, France; Laboratory of Cellular and Molecular Mechanisms of Hematological Disorders and Therapeutical Implications, Institut Imagine, Université Paris Cité, Inserm U1163, Paris, France; Initiatives IdEx Globule Rouge d’Excellence (InIdex GR-Ex), Université Paris Cité, Paris, France; Department of Internal Medicine, Reference Center for Sickle Cell Disease, Georges-Pompidou European Hospital, AP-HP, Université Paris Cité, Paris, France; Department of Internal Medicine, Reference Center for Sickle Cell Disease Tenon Hospital, AP-HP, Sorbonne University, Paris, France; Department of Biochemistry and Molecular Biology, Inserm UMR942, Lariboisière Hospital, AP-HP, Université Paris Cité, Paris, France; Hematology Department, Reference Center for Sickle Cell Disease, Robert Debré Hospital, AP-HP, Université Paris Cité, Paris, France; Pediatric Intensive Care Unit, Necker-Enfants Malades Hospital, AP-HP, Université Paris Cité, Paris, France; Department of Hematology, Necker-Enfants Malades Hospital, AP-HP, Université Paris Cité, Paris, France; Reference Center for Mastocytosis, Necker-Enfants Malades Hospital, AP-HP, Paris, France

## Abstract

A role for substance P in promoting neurogenic inflammation and pain has been described in sickle cell disease (SCD). However its origin and contribution to SCD pathophysiology remain unclear. We measured substance P level in plasma from 225 patients with SCD and observed the highest concentrations during acute chest syndrome (ACS). Therefore, we tested the hypothesis that substance P may induce ACS. In transgenic sickle mice, unlike control mice, intravenous injection of substance P caused lethal crises with dose-dependent acute lung injuries. Activation of FcεR1α with MAR-1 had similar effects, suggesting a role for mast cell or basophil activation and degranulation. Pretreatment of sickle mice with cromolyn, a stabilizer of mast cells and basophils, prevented lethal crisis and lung injuries induced by substance P injection. In SCD patients, blood cellular histamine levels and increased histidine decarboxylase activity were consistent with an involvement of circulating basophils. Flow cytometry analysis revealed higher basophil counts with increased activation and degranulation markers in patients compared with healthy controls. During vaso-occlusive crisis, absolute basophil counts tended to decrease, suggesting their recruitment outside the vascular compartment. The same results were observed in sickle mice after hypoxia-reoxygenation, intravenous hemin injection or substance P injection. Immunohistochemistry revealed the presence of mast cells and basophils in the lungs of sickle mice, but not in control mice, with further basophil recruitment and degranulation after intravenous substance P injection. In SCD patients, we observed extremely high levels of substance P in the sputum collected during ACS, consistently with mast cell and basophil degranulation in the lungs. In vitro, substance P was shown to be a potent chemoattractant for basophils via NK1R. Gene expression analysis on sorted circulating basophils from SCD patients revealed an increased expression of several chemokine receptors, including CCR3 and FPR1, which was confirmed by spectral flow cytometry and could contribute to the recruitment of basophils in the lungs. The two substance P receptors, NK1R and MRGPRX2, were also overexpressed, promoting the vicious cycle of substance P release and pain in SCD patients. Our results reveal a novel mechanism that contributes to the understanding of ACS pathogenesis and highlights the potential role of mast cells and basophils in SCD pathophysiology.

## Introduction

Sickle cell disease (SCD) is a severe hemoglobin disorder considered the most common monogenic disease worldwide with an incidence of 500 000 newborns affected per year.^1^ However our understanding of the complex multicellular pathophysiology of SCD is still limited. A role for mast cells has been suggested as high plasma levels of substance P and histamine have been reported in SCD patients at steady state with a further increase during vaso-occlusive crisis (VOC).^2–4^ In a transgenic sickle mouse model, the release of substance P by mast cells promoted nociceptor activation and neurogenic inflammation, characterized by increased microvascular permeability.^5^ By activating mast cells and enhancing the release of substance P, morphine paradoxically contributed to hyperalgesia and neuro-inflammation. An association between morphine and acute chest syndrome (ACS), a common form of acute lung injury in SCD, has long been suspected.^6^ ACS is a severe complication and a leading cause of admission to intensive care unit and premature death in SCD but its pathogenesis remains incompletely understood.^7,8^ Since lung injury during ACS is mainly characterized by edema formation, a role for substance P-induced plasma extravasation may be suspected.^9^ To our knowledge, the potential role of basophils has never been explored in SCD, although these innate immune cells share many morphological and functional similarities with mast cells, including ability to secrete histamine and substance P.^10,11^ Here, we investigate the hypothesis that substance P, released by mast cells and basophils, may be involved in ACS pathogenesis.

## Results

### Substance P is elevated in the plasma of SCD patients during ACS and in sickle mice after hypoxia/reoxygenation or hemin injection

In order to assess plasma substance P levels in SCD patients during ACS, we performed a prospective observational study in two French university-hospital SCD reference centers. We recruited 225 SCD patients and collected blood samples at steady state (n=132), during hospitalization for VOC without ACS (n=71) or during ACS (n=30) (Supplementary Table 1). Plasma Substance P level was higher than the upper limit of normal (ULN = 0.3 µg/L) in 4.5% of patients at steady state (n=6/132), in 85.9% during VOC (n=61/71) and in 100% during ACS (n = 30/30) (Figure 1A). Median [IQR] plasma substance P level was significantly higher during ACS than during VOC (1.11 [0.73-2.23] vs 0.73 [0.45-1.35] µg/L, p = 0.006). Among patients hospitalized for VOC, median substance P level was significantly higher in those receiving morphine than in those not receiving it (0.8 [0.52-1.47] vs 0.48 [0.38-0.76] µg/L, p = 0.039).

**Figure 1.**
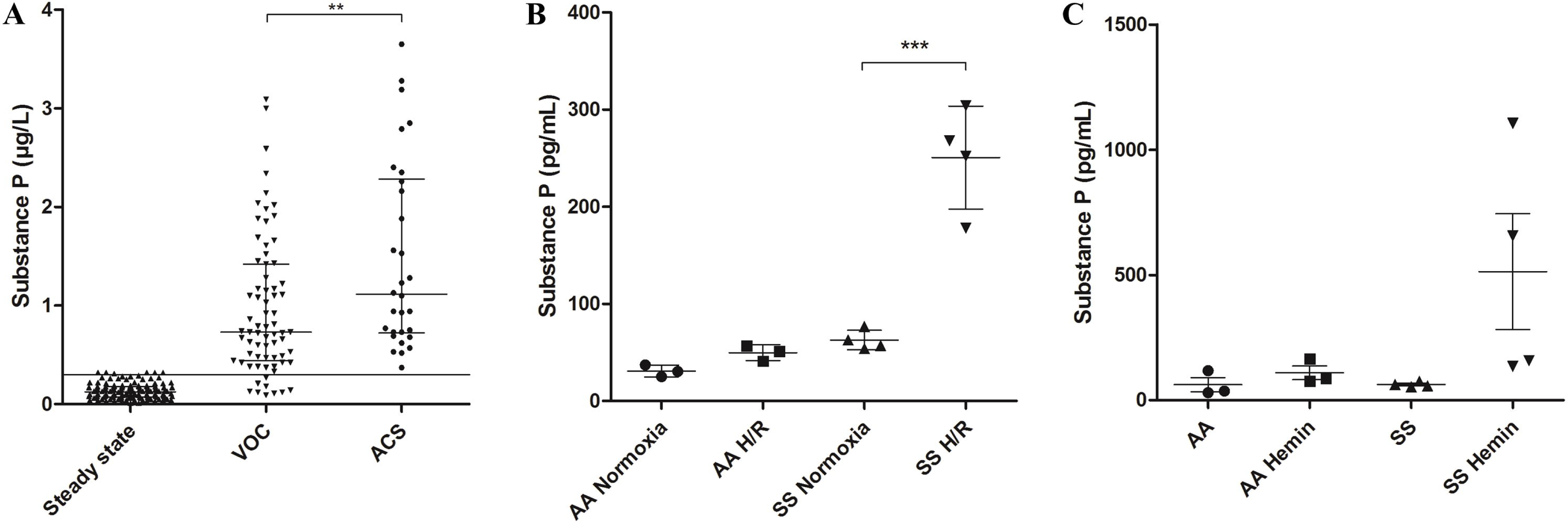
Plasma substance P levels are elevated during vaso-occlusive crisis (VOC) and acute chest syndrome (ACS) in patients with sickle cell disease (SCD) and SCD mice. (A) Plasma substance P levels in SCD patients were higher during hospitalization for VOC (n=71) than at steady sate (n=132) with further increase during ACS (n=30). Horizontal line is the 99^th^ percentile in the general population. (B-C) Plasma substance P levels were increased in SS mice but not in AA mice after hypoxia/reoxygenation (H/R) or hemin injection (70 µmol/kg i.v.). \*\*\**P*<0.001. \*\**P*<0.01.

To induce VOC or ACS, Townes transgenic sickle mice (herein referred as SS mice) and wild type human hemoglobin control mice (herein referred as AA mice) were exposed to hypoxia/reogygenation (8% oxygen for 3 hours followed by 21% oxygen for 1 hour), or they were infused with hemin (70 µmol/kg i.v.). In both cases, plasma substance P levels increased in SS but not in AA mice (Figure 1B and 1C). Similar to SCD patients, the highest plasma substance P levels were observed in SS mice that developed acute lung injuries.

### Substance P causes dose-dependent lethal crises and acute lung injuries in sickle mice

To test the hypothesis that substance P may induce VOC and ACS in sickle mice, SS and AA mice were infused with substance P (40mg/kg i.v.). Unlike AA mice, almost all SS mice developed intense pain, prostration and labored breathing leading to death within 3 hours (Figure 2A). Lethality rate was positively correlated with the dose of substance P (Figure 2B). Histological evaluation of the lungs of SS mice after substance P injection, revealed the presence of acute lung injuries, including edema, congestion, alveolar wall thickening and hemorrhage (Figure 2C). The lung injury score was significantly higher in SS mice than in AA mice, with further increase after substance P injection (Figure 2D). No injury was observed in the lungs of AA mice after i.v. substance P injection.

**Figure 2.**
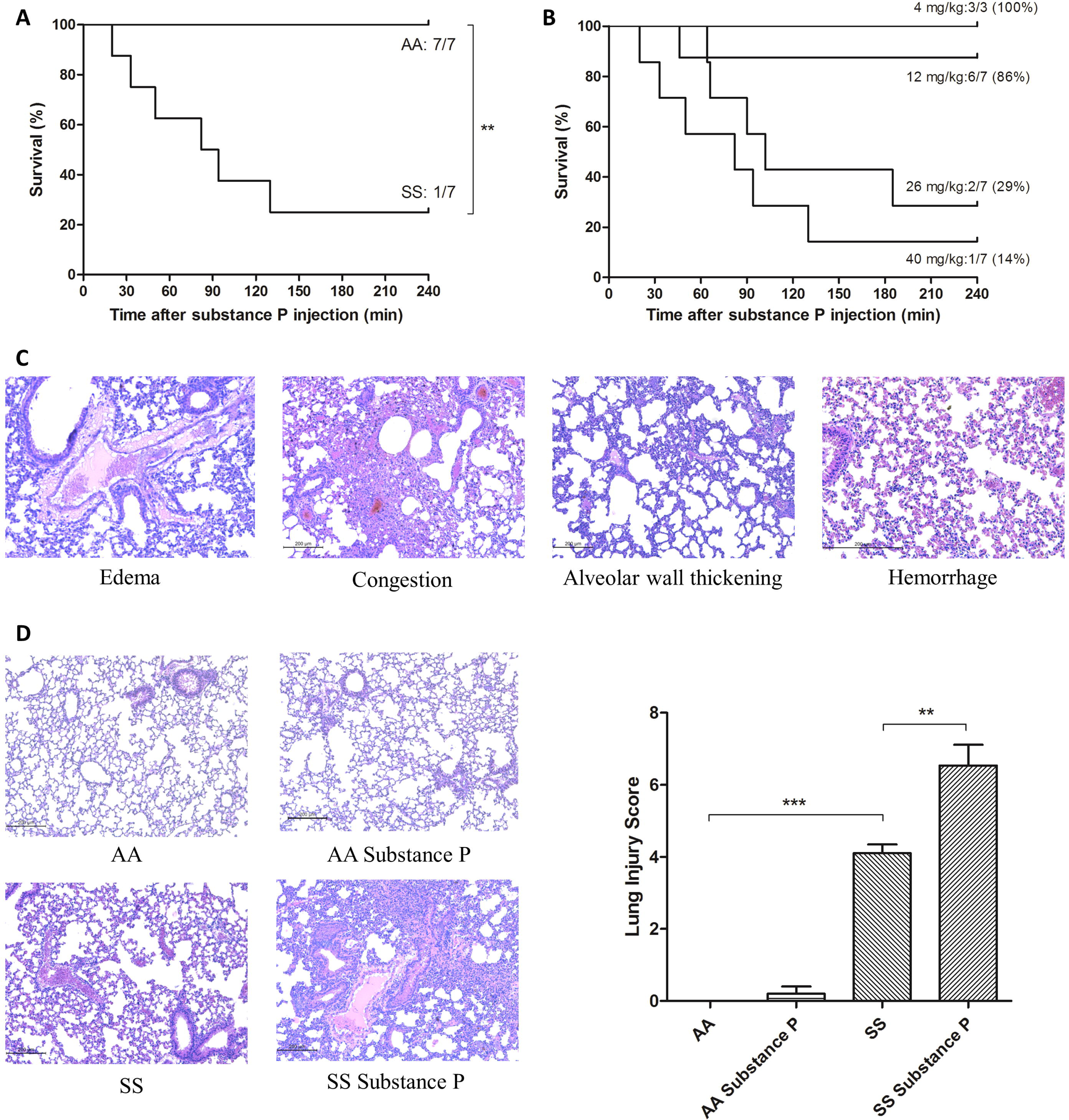
Substance P is responsible for lethal crises and acute lung injuries in SS mice. (A) Substance P injection (40 mg/kg i.v.) caused painful crises in all SS mice (n=7) but no AA mice (n=7), leading to death in the majority of SS mice (n=6/7). (B) Lethality induced by substance P was dose dependent. (C) Substance P injection was responsible for acute lung injuries with congestion, edema, alveolar wall thickening and hemorrhage. (D) Lung injury score was significantly higher in SS mice than in AA mice, with further increase after substance P injection. \*\*\**P*<0.001. \*\**P*<0.01.

### Activation of Fc_ε_R1_α_ causes lethal crisis and lung injuries in sickle mice, whereas cromolyn prevents VOC and lung injuries induced by substance P, which suggests a role for mast cells and/or basophils

It has been previously reported in sickle mice that substance P released from sensory nerve endings and from mast cells can in turn induce mast cell activation and degranulation, thus contributing to a vicious cycle of substance P release.^5^ The same auto-amplification loop may be observed with basophils, as they also express the two substance P receptors, NK1R and MRGPRX2 (Mrgprb2 in mice). To test the hypothesis that activation and degranulation of mast cells and basophils may contribute to VOC/ACS, SS and AA mice received i.p. injection of MAR-1 (10 µg/kg), an anti-FcεR1α monoclonal antibody that induces mast cell/basophil activation. Unlike AA mice, all SS mice developed intense pain and the large majority died within 3 hours (Figure 3A). Histological evaluation of their lungs revealed the presence of edema, congestion, alveolar wall thickening and hemorrhage (Figure 3B). Subsequently, to investigate if substance P induces VOC/ACS through mast cell/basophil degranulation, SS mice were pretreated with cromolyn (10 mg/kg/day i.p.) for four days before substance P injection (12 mg/kg i.v.) in order to inhibit mast cell/basophil degranulation. No adverse effects were seen in cromolyn-pretreated SS mice, whereas all non-pretreated SS mice developed intense pain and prostration in the first 30 minutes after substance P injection, leading to death after 45 minutes in one out of four mice. Furthermore, lung histology of cromolyn-pretreated SS mice showed significantly reduced lung injury after substance P injection compared to non-pretreated SS mice (Figure 3C and 3D). Together, these results support the hypothesis that substance P-mediated activation and degranulation of mast cells and/or basophils, promote VOC and ACS.

**Figure 3.**
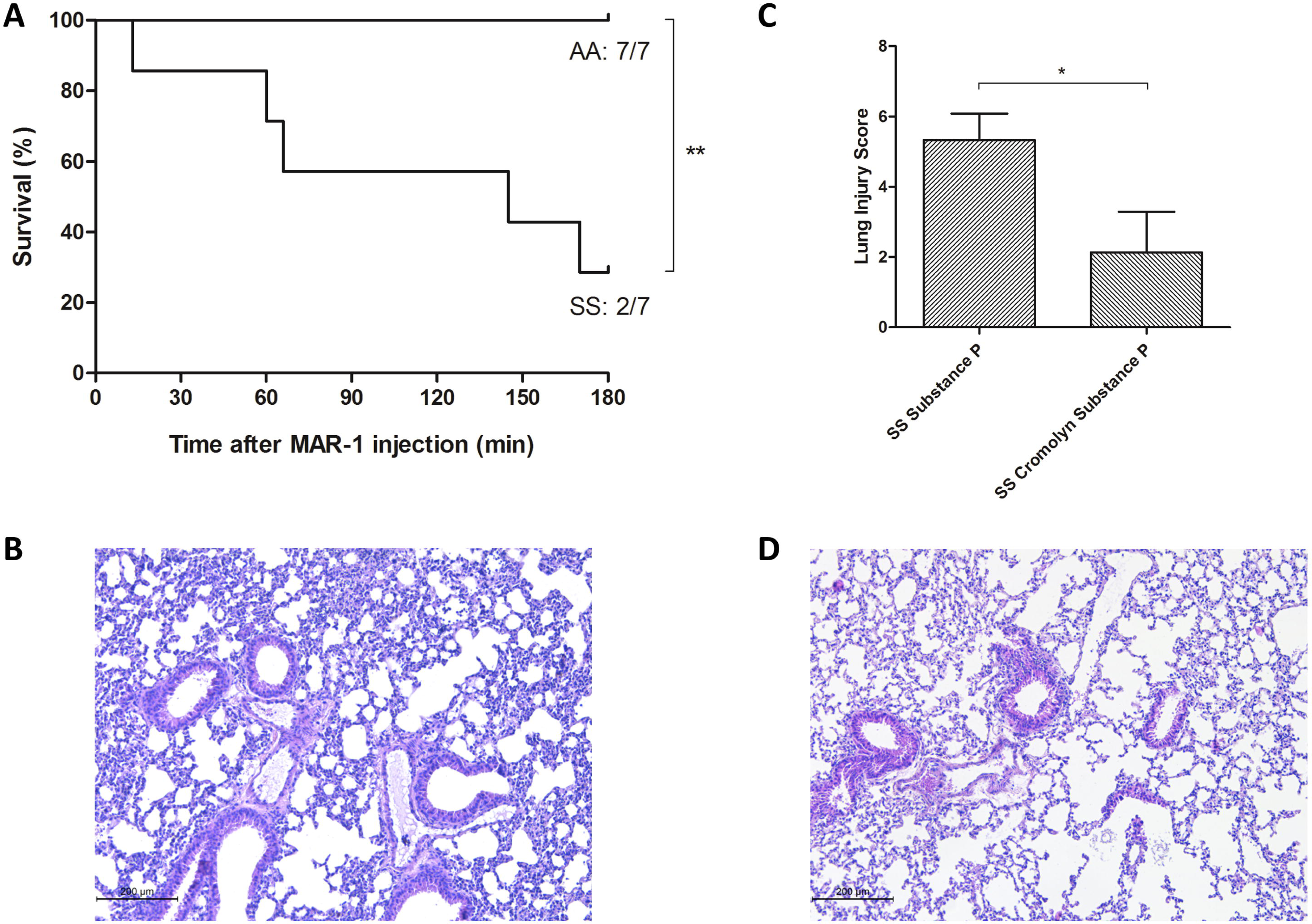
**MAR-1 (anti-Fc**_ε_**R1**_α_**) induces lethal crises and acute lung injuries in SS mice whereas cromolyn prevents lung injuries induced by substance P.** (A) MAR-1 injection (10 µg/kg i.p.) caused painful crises in all SS mice (n=7) but no AA mice (n=7), leading to death in the majority of SS mice (n=5/7). (B) Lung injuries induced by MAR-1 were similar to those induced by substance P, with increased congestion, edema, alveolar wall thickening and hemorrhage. (C-D) Lung injuries induced by substance P (12 mg/kg i.v.) in SS mice (n=4) were significantly reduced by pretreatment with cromolyn (10 mg/kg/d i.p., n=4) for four days. \*\**P*<0.01. \**P*<0.05.

### Basophils display an activated phenotype and are more numerous in the blood from SCD patients at steady state than in controls, while their number tends to decrease during VOC

In order to assess the number and activation/degranulation status of circulating basophils by flow cytometry, blood samples from SCD patients were collected (1) at steady state (n=8), (2) just before transfusion during a monthly exchange transfusion (MET) program (n=15), or (3) during a VOC (n=10), and were compared to age-matched controls (n=15). The number of basophils was significantly increased in off-crisis patients (MET and steady state) compared to controls, while it tended to decrease during VOC, suggesting their recruitment outside the vascular compartment (Figure 4A). The expression of the activation marker CD203c was significantly increased on basophils from off-crisis SCD patients, compared to controls (Figure 4B). The expression of the degranulation marker CD63 was also significantly increased in patients at steady state compared to controls (Figure 4C).

**Figure 4.**
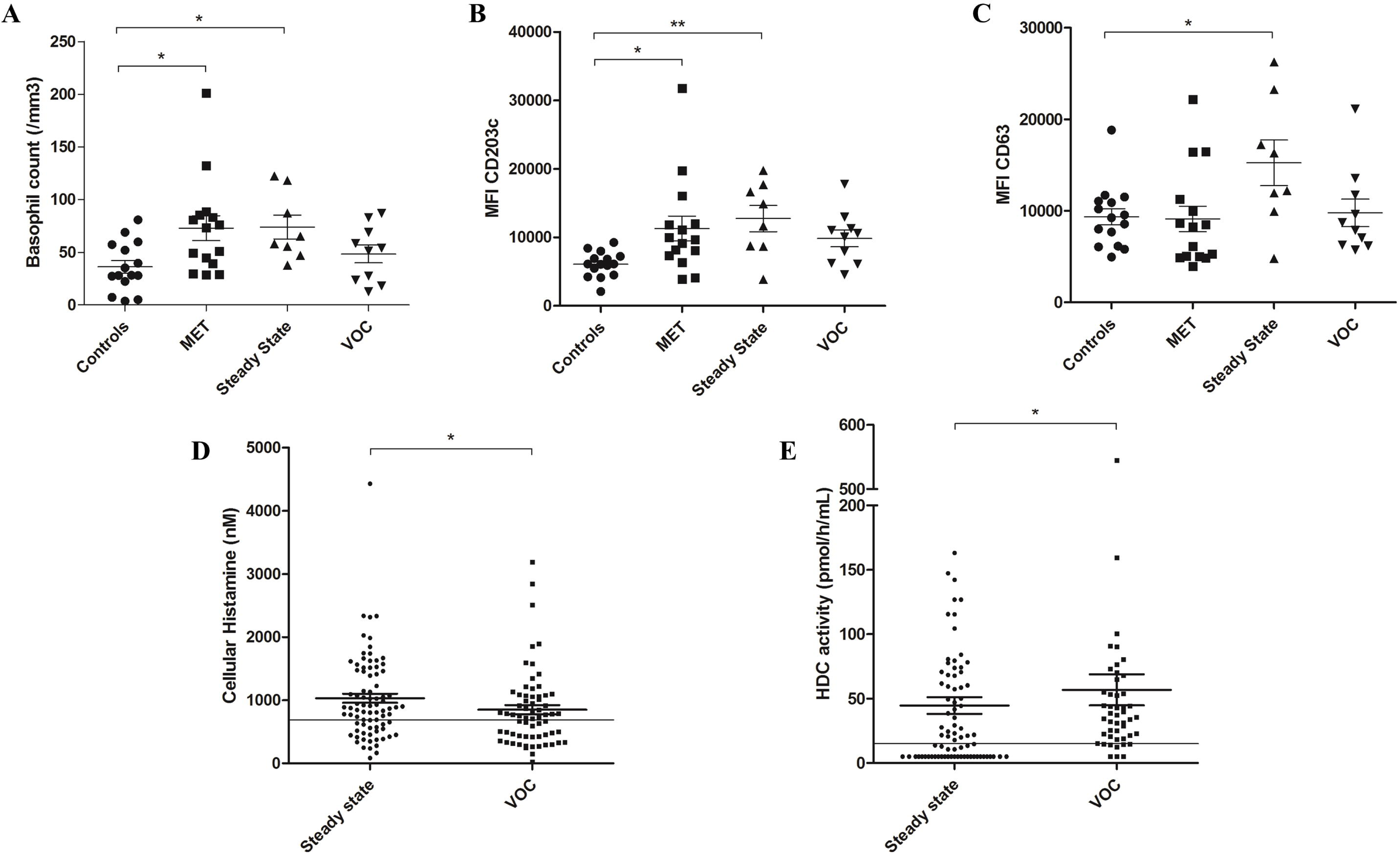
Basophils from SCD patients are more numerous and show increased activation/degranulation. (A) Basophil counts by flow cytometry were increased in SCD patients at steady state (n=8) and during a monthly exchange transfusion (MET) program (n=15), but not during VOC (n=10), compared to age-matched controls. (B) The expression of the activation marker CD203c was increased on basophils from SCD patients at steady state and during a MET, compared to controls (C). The expression of the degranulation marker CD63 was increased in patients at steady state compared to controls. (D) Cellular histamine level was higher than ULN in 65.9% of patients at steady state (n=54/82) and in 56.1% during VOC (n=37/66). (E) Total blood histidine decarboxylase (HDC) activity was higher than ULN in 55% of patients at steady state (n=44/80) and in 83.3% during VOC (n=40/48), reflecting basophil activation with increased histamine production at steady state and even more during VOC. \*\**P*<0.01. \**P*<0.05.

We have previously reported high levels of plasma histamine in a large cohort of SCD patients with further increase during VOC.^4^ In order to investigate the possible involvement of circulating basophils, we measured plasma and total blood histamine levels, allowing to calculate cellular histamine levels, in 148 SCD patients at steady state (n=82) and during VOC (n=66). Cellular histamine levels were higher than ULN in 65.9% of patients at steady state (54/82) and in 56.1% during VOC (37/66) (Figure 4D). Since mast cells are classically not circulating blood cells, it probably reflects the increased number of circulating basophils, as well as increased histamine production due to basophil activation. This hypothesis was further supported by total blood histidine decarboxylase (HDC) activity, which was found higher than ULN in 55% of patients at steady state (44/80) and in 83.3% during VOC (40/48), suggesting basophil activation with increased histamine production at steady state and even more during VOC (Figure 4E). The decrease in cellular histamine levels observed during VOC may reflect increased degranulation or recruitment of basophils outside the vascular compartment.

### Basophils and mast cells from sickle mice are recruited to the lungs

Similar to what we observed in SCD patients, blood basophil counts in SS mice were significantly higher than in AA mice and tended to decrease during VOC/ACS induced by hypoxia/reoxygenation, hemin or substance P (Figure 5A, 5B and 5C). In order to investigate if circulating basophils could be recruited to the lungs, thereby contributing to observed injuries, fluorescent immunohistochemistry was performed on the lungs of SS and AA mice that were injected with substance P (12 mg/kg i.v.). We found that FcεR1α^+^ cells were present at steady state in the lungs of SS mice but not AA mice and were more numerous after substance P injection in SS mice (Figure 5D). This effect was abrogated in cromolyn-pretreated mice, suggesting a role for mast cell/basophil degranulation in promoting basophil recruitment to the lungs. Subsequently, lung immunohistochemistry was performed with antibodies directed against FcεR1α, c-kit and Mcpt8, in order to better distinguish mast cells and basophils. We found an unexpectedly high number of both mast cells and basophils in the lungs of SS mice at steady state and during VOC/ACS, with a possibly enhanced degranulation after substance P injection. By contrast, no mast cells and no basophils were found in the lungs of AA mice (Figure 6A). In order to assess the potential effects of substance P on mast cell/basophil degranulation, murine bone marrow-derived mast cells and basophils were incubated with avidin, with or without substance P, for 30 minutes and the percentage of avidin+ cells was measured by flow cytometry. Substance P was responsible for a significant increase in avidin+ cells, reflecting increased degranulation (Figure 6B-C). Thus, our results reveal the presence of mast cells and basophils in the lungs of SS mice and suggest a role for their degranulation in ACS pathogenesis.

**Figure 5.**
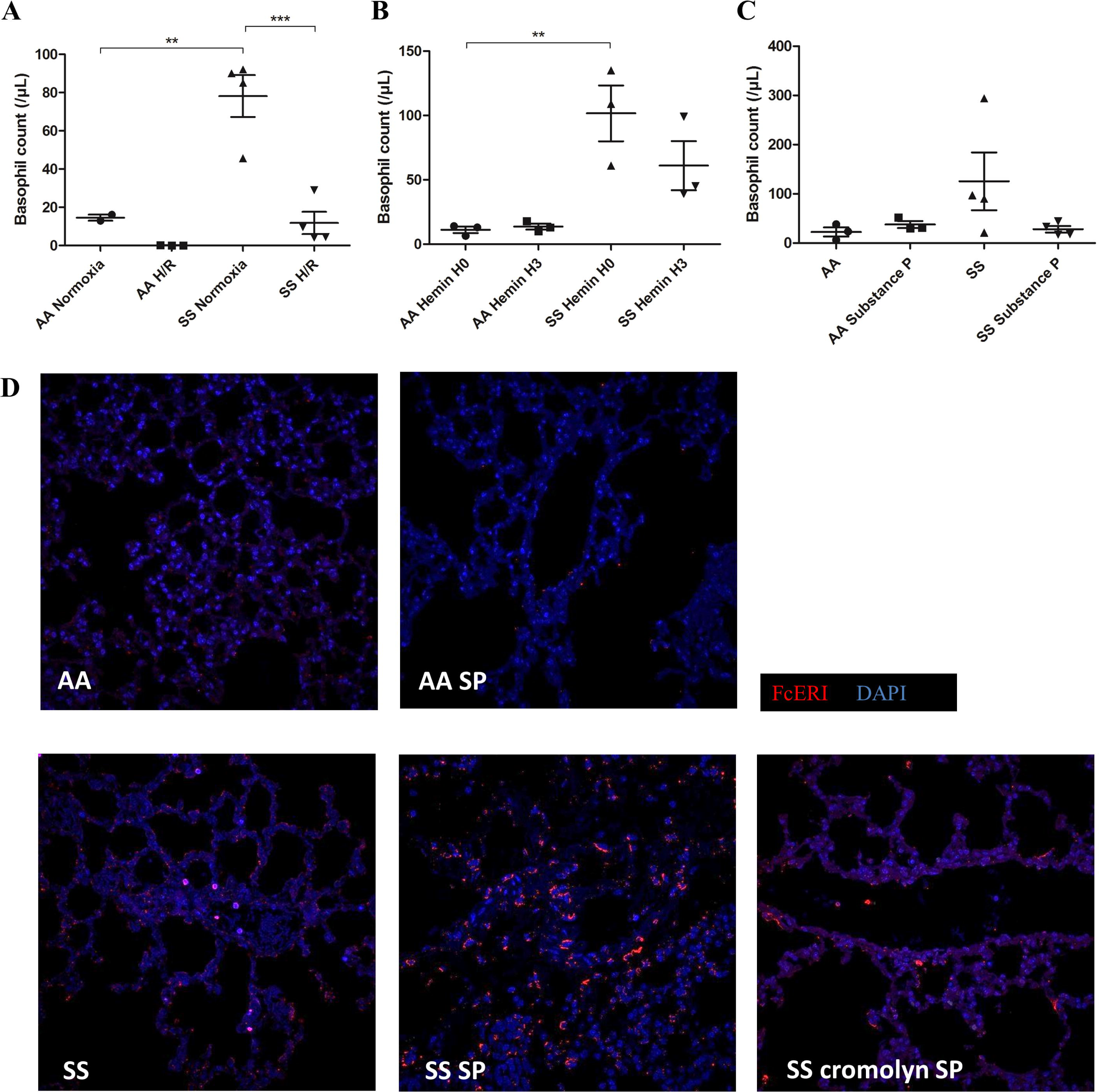
Basophils from SCD mice are recruited to the lungs during VOC/ACS. (A-C) Blood basophil counts from SS mice were higher compared to AA mice and decreased during VOC/ACS induced by H/R, hemin or substance P. (D) fluorescent immunohistochemistry on the lungs of SS and AA mice injected with substance P (12 mg/kg i.v.) revealed the presence of FcεR1α ^+^cells (stained in red; nuclei stained in blue by DAPI) at baseline in the lungs of SS mice but not AA mice, with further increase after substance P injection in SS mice. This effect was abrogated in mice that were pretreated with cromolyn, suggesting a role for mast cell/basophil degranulation in promoting basophil recruitment to the lungs. \*\*\**P*<0.001. \*\**P*<0.01.

**Figure 6.**
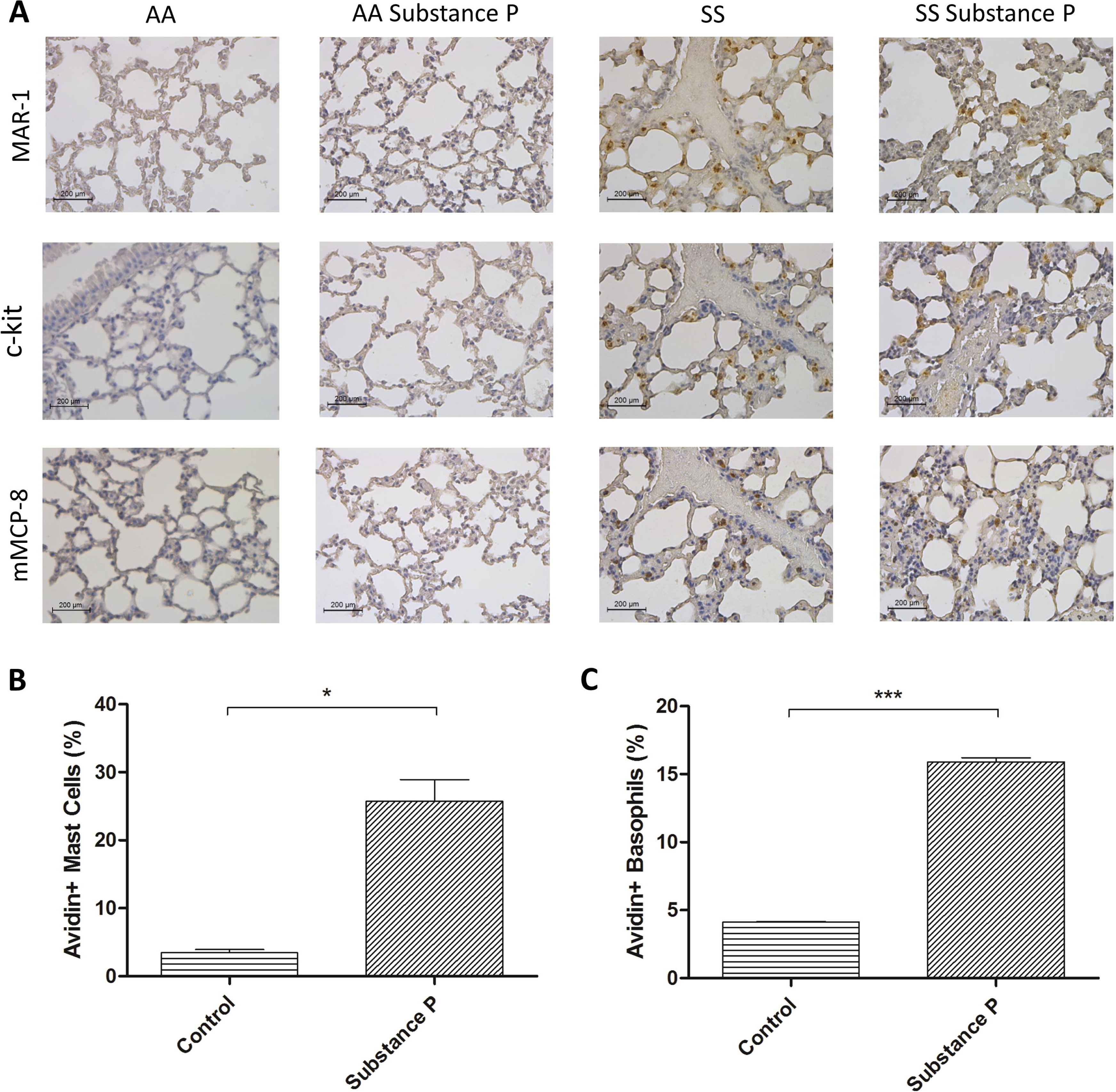
Mast cells and basophils are present in the lungs of SS mice but not AA mice and substance P induces mast cell/basophil degranulation. (A) Immunohistochemistry revealed a high number of both mast cells (stained by anti-FcεRIα (MAR-1) and anti-c-kit primary antibodies) and basophils (stained by anti-FcεRIα (MAR-1) and anti-Mcpt-8 primary antibodies) in the lungs of SS mice, but not in AA mice. (B-C) In vitro, incubation of murine bone marrow derived mast cells (B) and basophils (C) with avidin (5µg/ml) and substance P (10^-6^ M) was responsible for a significant increase in avidin+ cells measured by flow cytometry, reflecting increased degranulation. \*\*\**P*<0.001. \**P*<0.05.

### Substance P levels are dramatically high in the sputum from SCD patients during ACS, and substance P is a potent chemoattractant for basophils in vitro

Because immunohistochemistry of human SCD lungs was not possible, we decided to explore mast cell/basophil degranulation in the lungs by measuring substance P and histamine levels in the sputum obtained from SCD patients during ACS (n=7), compared with sputum from SCD patients (n=4) and age-matched controls (n=6) affected by viral or bacterial pneumonia. Substance P levels measured by radioimmunoassay were dramatically increased (30-fold) in the golden sputum from SCD patients during ACS (Figure 7A). Histamine levels were also increased but to a much lesser degree, which suggests a prominent role for substance P in ACS pathogenesis (Figure 7B). It has been previously reported that substance P may have a chemoattractant effect on human basophils via NK1R.^12^ To confirm this finding, we studied the migration of murine bone marrow-derived basophils in vitro, using microchemotaxis chambers. Substance P chemoattractant effect was found as potent as that of LPS and was abrogated by NK1R antagonist (Figure 7C). Histamine was also confirmed to be a potent chemoattractant for basophils.

**Figure 7.**
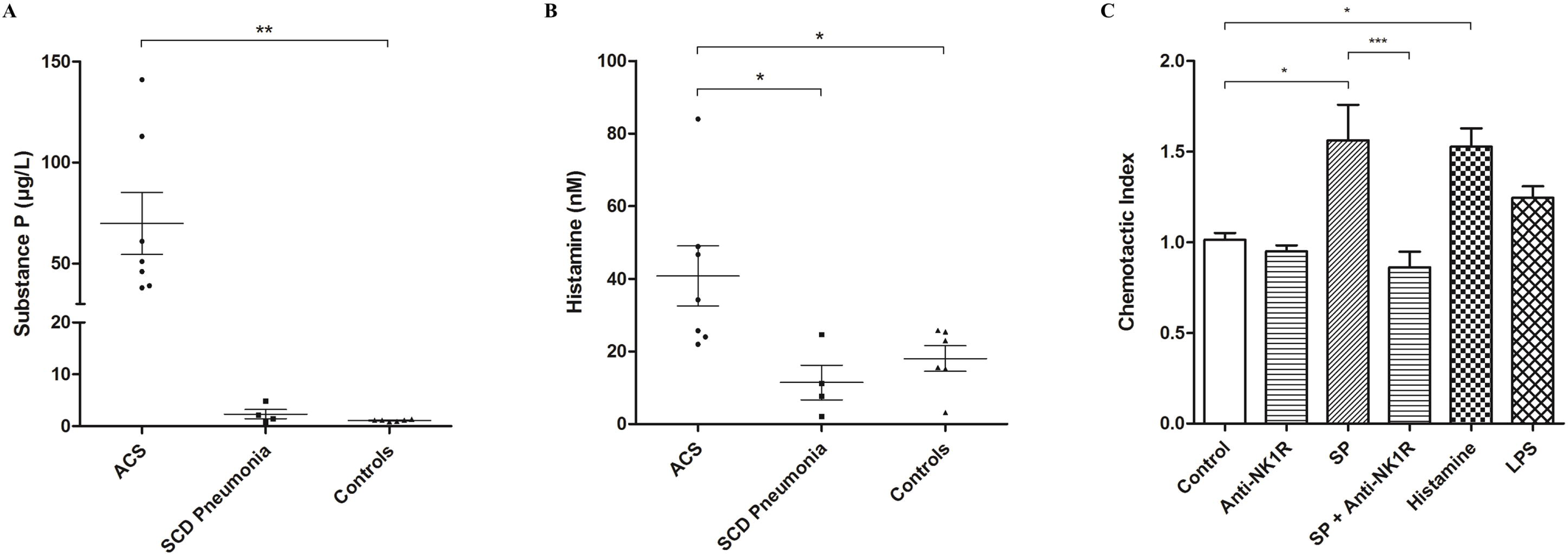
Substance P levels are extremely high in the sputum from SCD patients during ACS and substance P has a potent chemoattractant effect on basophils. (A-B) Substance P levels, and to a lesser degree histamine levels, were higher in the sputum from SCD patients during ACS (n=7) than in the sputum from SCD patients (n=4) and age-matched controls (n=6) during pneumonia. (C) In vitro, substance P had a chemoattractant effect on murine bone marrow-derived basophils, which was abrogated by NK1R antagonist. LPS was used as a positive control and histamine was also confirmed to be a potent chemoattractant for basophils. \*\*\**P*<0.001. \*\**P*<0.01. \**P*<0.05.

### Basophils from SCD patients overexpress several chemokine receptors and the two substance P receptors, NK1R and MRGPRX2

To further investigate the mechanisms underlying basophil recruitment to the lungs, blood basophils from SCD patients (at steady state (n=5), during a MET program (n=5), during a VOC (n=5), or at the end of a VOC (n=5)) and age-matched healthy controls (n=5) were isolated by flow cytometry sorting, allowing for gene expression analysis of several chemokine receptors. Using the “human chemokines and receptors RT² Profiler PCR Array”, we found an overexpression of CCR2, CCR3 and FPR1 chemokine receptors (Figure 8A, 8B and 8C). To confirm that gene overexpression was associated with enhanced expression of these chemokine receptors on the surface of basophils, spectral flow cytometry was performed on blood samples from off-crisis SCD patients (n=10) and age-matched healthy controls (n=5). CCR2 expression was not significantly increased in SCD patients (Figure 8D) but we found a significant overexpression of CCR3 (Figure 8E) and FPR1 (Figure 8F) compared to controls. These results suggest that activated basophils from SCD patients are prone to respond to various chemokines via CCR3, as well as to N-formylmethionine-containing oligopeptides, released by pathogens and injured tissues, via FPR1. Spectral flow cytometry analysis of circulating basophils from SCD patients also revealed significantly increased expression of the two main substance P receptors, NK1R and MRGPRX2, compared to basophils from healthy controls (Figure 8G and Figure 8H). Hence, activated basophils from SCD patients may be more prone to respond to substance P and morphine, thereby contributing to the vicious cycle of inflammation and pain mediated by substance P. As previously observed, basophil count was significantly higher in SCD patients at steady state than in controls (Figure 8I).

**Figure 8.**
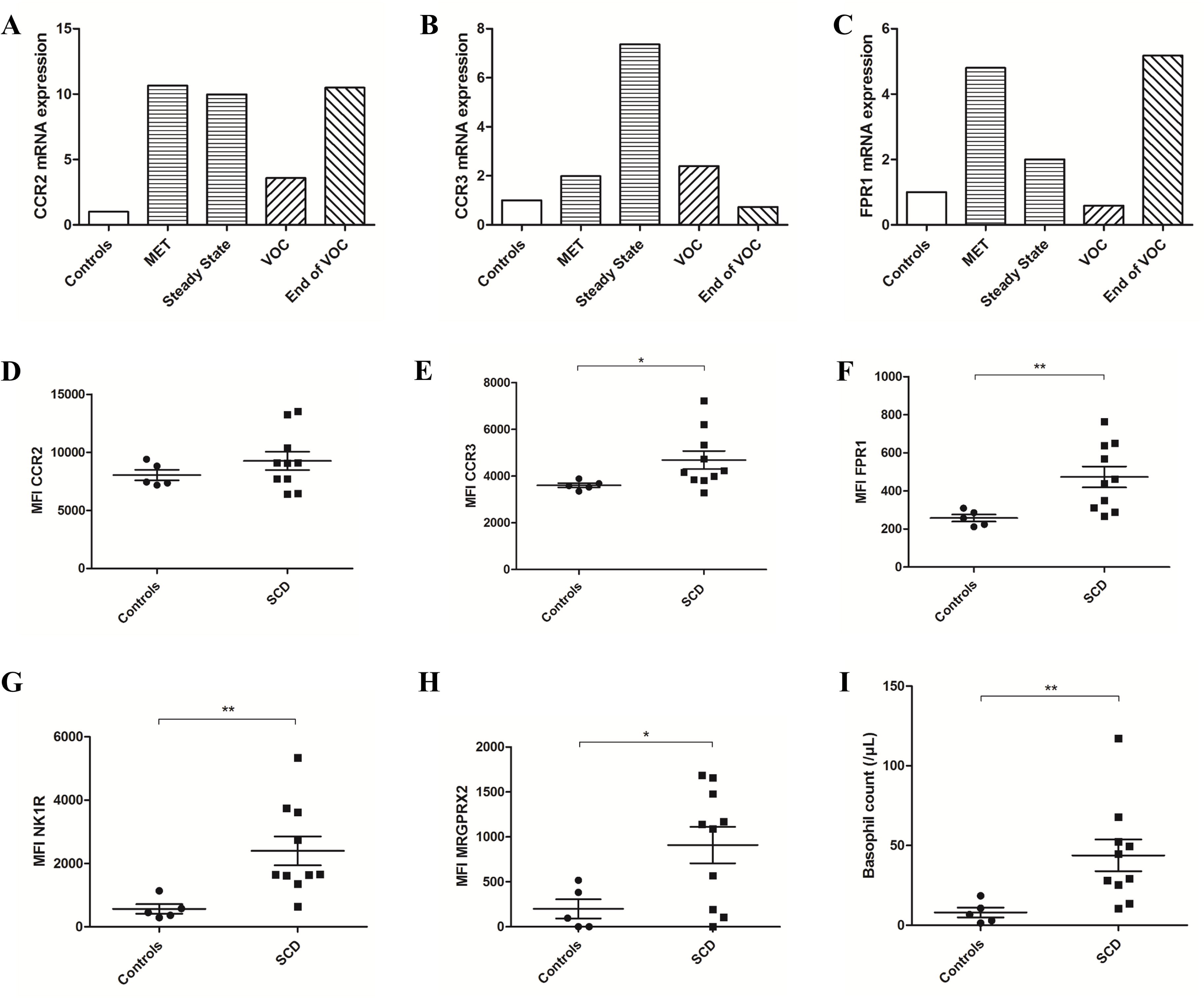
Basophils from SCD patients overexpress several chemokine receptors and the two substance P receptors, NK1R and MRGPRX2. (A-C) Gene expression of CCR2, CCR3 and FPR1 was increased in blood basophils from SCD patients isolated by flow cytometry sorting (at steady state (n=5), during a MET program (n=5), during a VOC (n=5) or at the end of a VOC (n=5)) compared to age-matched healthy controls (n=5). (D-F) Subsequently, spectral flow cytometry on blood samples from off-crisis SCD patients (n=10) and age-matched healthy controls (n=5) revealed increased basophil surface expression of CCR3 and FPR1 compared to controls. (G-H) The two substance P receptors, NK1R and MRGPRX2 were also overexpressed. (I) Basophil count was higher in SCD patients than in controls. \*\**P*<0.01. \**P*<0.05.

Together, our results suggest a role for basophils in SCD pathophysiology and reveal a novel mechanism involving activated basophils, mast cells and substance P in ACS pathogenesis.

## Discussion

Here, we report for the first time an involvement of basophils in SCD pathophysiology and a potential role for substance P released by mast cells and basophils in ACS pathogenesis. First, we found that substance P levels were extremely high in plasma from SCD patients during ACS and that substance P injection to SS mice, unlike AA mice, was responsible for lethal crises with acute lung injuries, mediated by mast cell/basophil degranulation. Second, we showed that basophils from SCD patients are activated and that their number is increased at steady state compared to basophils from healthy controls, while it decreases during VOC/ACS, which suggests recruitment outside the vascular compartment. Third, we found an unexpectedly high number of mast cells and basophils in the lungs from SS mice at steady state and during VOC/ACS, with enhanced degranulation after substance P injection. Consistently with these results, substance P levels were dramatically high in the sputum from SCD patients during ACS, which probably reflects an auto-amplification loop, mediated by activated mast cells and basophils. Fourth, we demonstrated that basophil recruitment to the lungs may be promoted by the chemoattractant effect of substance P via NK1R, whose expression is increased on blood basophils from SCD patients. MRGPRX2, the other receptor for substance P and for morphine, was also overexpressed, as where the chemokine receptors CCR3 and FPR1. Finally, we demonstrated the prophylactic role of targeting mast cell/basophil degranulation with cromolyn in SCD mice, thus opening new therapeutic perspectives in SCD management.

During the past decade, growing evidence has suggested a role for mast cells in SCD pathophysiology. In SCD mice, mast cell activation was found responsible for increased levels of tryptase and substance P in the skin, as well as increased release of substance P in dorsal root ganglion, thus promoting hyperalgesia and neurogenic inflammation.^5,13^ Inhibition of mast cells with the *c-kit*/tyrosine kinase inhibitor imatinib or with cromolyn decreased tryptase and substance P levels, as well as neurogenic inflammation and hyperalgesia.^5^ In vitro, activated mast cells from SCD mice were also shown to promote endoplasmic reticulum stress in brain endothelial cells, resulting in upregulated P-selectin expression with increased endothelial permeability.^14^ Hence, mast cells may be involved in SCD blood brain barrier dysfunction. In SCD patients, high plasma levels of both histamine and substance P have been reported, which suggests a release by mast cells. However, in a large cohort of SCD patients, plasma tryptase levels were not increased, which may be interpreted as an activation of mast cells rather than an increase in their number, but may also reflect the involvement of another circulating blood cell that produces histamine and substance P but not tryptase, namely the basophil.^4^ Here we report increased levels of both cellular histamine and HDC activity in the blood from SCD patients, which supports our hypothesis since mast cells, unlike basophils, are not circulating blood cells.^15^

Basophils are potent effectors of the innate immune system, sharing many structural and functional similarities with mast cells.^16^ Because they are one of the less numerous leukocyte populations, basophils have long been neglected, apart from their role in allergy and defense against parasitic infections.^17^ However in the last years, several studies have highlighted their powerful immune regulatory functions, as well as deleterious contribution to autoinflammatory and autoimmune diseases.^18–20^ Because SCD is considered a chronic inflammatory disease with enhanced autoimmunity, an involvement of basophils could legitimately have been suspected.^21,22^ Here we show that basophils are more numerous and display enhanced activation and degranulation in SCD patients compared to healthy controls. Moreover, due to increased expression of the two substance P receptors, NK1R and MRGPRX2, basophils may be more prone to be activated by substance P, thereby promoting a vicious cycle of substance P release and mast cell/basophil activation. An association between MRGPRX2 surface expression on human basophils and their activation status has been previously evoked.^23^ Importantly, MRGPRX2 has also been shown to mediate mast cell degranulation in response to morphine and is likely to have the same effect on basophils.^24^ Therefore, morphine which is considered a reference analgesic in VOC management may paradoxically have deleterious effects by inducing the release of substance P by mast cells and basophils. Consistently with this hypothesis, we found that among patients hospitalized for VOC, plasma substance P level was significantly higher for those receiving morphine. Hence, our finding that substance P induces lethal crises with lung injuries in SCD mice suggests that the long-date suspected association between morphine and ACS may be the consequence of increased substance P release by mast cells and basophils in response to morphine.

Substance P is a neuropeptide that acts as a primary pain neurotransmitter and a neuromodulator, encoded by the *TAC1* gene in neurons as well as in several immune cells, including mast cells and basophils.^5,10,25^ Via NK1R, substance P contributes to edema and neurogenic inflammation by increasing venular permeability and plasma extravasation, but it also acts on mast cells and basophils themselves, thereby promoting a vicious cycle of cell activation and substance P release.^5,10^ Here, we report an elevation of plasma substance P levels in SCD patients during ACS compared to VOC, as well as dramatically increased substance P levels in the sputum from SCD patients during ACS. Therefore, substance P could be used as a specific marker of ACS, especially in some cases where the distinction between ACS and pneumonia is difficult. More importantly, these results may reflect a role for substance P in ACS pathogenesis, as strongly suggested by the observation of acute lung injuries in SS mice, but not AA mice, after substance P injection. Similar effects of the anti-FcεR1α monoclonal antibody and prevention of substance P-induced acute lung injuries by cromolyn further support the hypothesis that substance P deleterious effects are mediated by the degranulation of mast cells and basophils observed in the lungs from SS mice, unlike AA mice.

The mechanisms underlying the recruitment of mast cells and basophils to the lungs from SS mice, but not AA mice, remain to be determined. Gene expression analysis and flow cytometry on circulating basophils from SCD patients revealed upregulated expression of several chemokine receptors, including CCR3 and FPR1. Hence, activated basophils from SCD patients may be recruited to the lungs by chemokines or by N-formylmethionine-containing oligopeptides released by pathogens and injured pulmonary tissues. In the lungs of SCD mice, histological signs of inflammation and increased levels of cytokines and chemokines have been reported.^26^ Similarly, we observed increased levels of several chemokines, including CCL2 and CCL3, in the sputum from SCD patients.^27^ Moreover, we have shown that substance P acts as a potent chemoattractant for basophils via NK1R, whose expression is increased on their surface in SCD patients. Of note, histamine was also confirmed to have a powerful chemoattractant effect on basophils, and we found increased levels of both substance P and histamine in the sputum from SCD patients during ACS.

Regarding the suspected mechanisms of basophil activation in SCD, a role for hemoglobin S and heme released by chronic hemolysis may be suspected, by analogy to other innate immune cells, as basophils express TLR4.^28^ Consistently with this hypothesis, plasma substance P levels from SCD patients have been previously found positively correlated with hemolysis markers.^3^ Furthermore, we found that hemin injection to SCD mice was responsible for increased plasma substance P levels. On the other hand, plasma histamine levels from SCD patients were found positively correlated with C-reactive protein (CRP) levels, which suggests a possible role for inflammation in the mechanisms leading to mast cell/basophil activation.^4^ SCD can be considered a chronic inflammatory disease, with increased levels of various cytokines, and both mast cells and basophils possess a large repertoire of cytokine receptors, enabling them to respond to inflammatory signals.^29–31^

A role for heme in inducing ACS in SCD mice has been previously described and was found mediated by endothelial P-selectin upregulation through TLR4 activation.^32–34^ This model is not inconsistent with our findings on substance P involvement in ACS since increased substance P plasma levels have been observed after hemin injection to SCD mice. Vascular stasis induced by heme-mediated endothelial P-selectin expression may contribute to mast cell/basophil activation and degranulation, ultimately resulting in increased substance P release. Indeed, mast cells can be activated by ischemia/reperfusion and are considered important actors of the post-ischemic inflammatory response.^35–37^ Our observation of increased plasma substance P and histamine levels after hypoxia/reoxygenation supports this hypothesis. Finally, an association between aeroallergen sensitization and ACS incidence has been reported, which is consistent with our hypothesis of a role for mast cells, basophils and substance P in ACS pathogenesis since allergy may contribute to ACS by promoting the release of mast cell/basophil mediators such as substance P.^38^

In conclusion, our findings reveal a mechanism that involves mast cells, basophils and substance P in the pathogenesis of ACS and open new therapeutic perspectives in SCD.

## Methods

### Patients

A prospective observational study was performed in two French university-hospital SCD reference centers. Eligibility criteria were SCD of all types including SS, SC, S/β^0^ and S/β^+^, and age ≥ 1 year. Exclusion criteria were other diseases resulting in high blood substance P and histamine levels or possibly modifying basophils (e.g., mastocytosis, symptomatic allergy, parasitic infections, inflammatory or autoimmune diseases such as systemic lupus erythematosus) and recent immunosuppressive or antihistamine treatment. All patients were recruited: 1) at steady state; 2) before transfusion during a MET program; 3) during hospitalization for VOC; 4) at the end of VOC just before discharge; 5) during hospitalization for ACS; and 6) during hospitalization for pneumonia. Controls were recruited among unaffected siblings of SCD patients. Blood was collected in ethylenediamine tetra-acetic acid (EDTA), and plasma obtained by centrifugation (10 min, 3,500 g, 4°C) was stored at −80°C. Sputum samples collected by chest physiotherapy during ACS or during pneumonia were immediately stored at −80°C. Several clinical and biological data were obtained from the patient medical records (Supplementary Table 1). Informed consent was obtained from adult patients and from parents or legal guardians for children (age < 18 years). The study was approved by a medical ethics committee (GR-Ex/CPP-DC2016-2618/CNIL-MR01).

### Mice and procedures

We have used male and female HbAA and HbSS-Townes transgenic sickle mice, obtained from colonies established in our laboratory.^39^ Mouse genotypes were assessed by PCR, using protocols established by the Jackson Laboratories (USA). All mice used in our experiments were 12 to 14-week-old and were pathogen-free. Mice were injected with freshly prepared solutions of hemin (70 µmol/kg i.v.), substance P (4 to 40 mg/kg i.v.) or MAR-1 (anti-FcεR1α; 10 µg/kg i.p.). Pretreatment with cromolyn was administered for four days (10 mg/kg/day i.p.) before substance P injection. For hypoxia/reoxygenation experiments, mice were exposed to 8% oxygen for 3 hours in a hypoxia chamber (Biospherix-ProOx P360), followed by reoxygenation for 1 hour in room air. Blood samples were obtained by retro-orbital sinus puncture with capillary tubes internally coated with heparin/EDTA anticoagulant. Lungs were processed immediately after death by cervical dislocation and were fixed in 10% buffered formalin (Sigma-Aldrich). All procedures on mice were performed in compliance with the local ethics committee. Ethical approval was obtained from the Institutional Animal Care and Use Committee (IACUC) and authorized by the French Ministry for Research (201809051638898).

### Reagents and drugs

Hemin [Ferric PPIX; Sigma-Aldrich] was prepared by dissolving in 0.25M NaOH, before pH neutralization with HCl and concentration adjustment with PBS. It was then filter sterilized before immediate use. Substance P (Tocris Bioscience) was prepared by dissolving in PBS and was used immediately. MAR-1 (eBioscience) and Cromolyn (Sigma) were prepared by dissolving in PBS.

### Substance P, histamine and HDC activity measurement

Plasma substance P and cellular histamine were measured by radioimmunoassay (Beckman-Coulter IM1659) and HDC activity was measured by fluorescent enzyme-linked immunoassay (UniCAP; Pharmacia/Thermo Fisher). In humans, the upper limit of normal (ULN) for plasma substance P (0.3 µg/L), blood cellular histamine (690 nM) and blood HDC activity (15 pmol/h/mL) were defined as the 99^th^ percentile in the general population.

### Histopathology

Lung tissue sections were stained with hematoxylin and eosin, before examination with a Leica DM2000 LED microscope. Images were recorded with a Leica MC170 HD camera and Leica Application Suite software (version 4.7.1). To score lung injury, we used a scoring system based on 4 parameters: edema, hemorrhage, congestion and alveolar wall thickening. For each parameter, the following notation was applied: 0, absent; 1, mild; 2, moderate; 3, severe; 4, very severe. Individual scores ranging from 0 to 16 were calculated in 5 random focal areas for each lung section (magnification, x100), allowing to calculate an overall lung injury score which was the mean of these 5 individual scores.

### Immunofluorescence and immunohistochemistry

Paraffin slides of lung tissue sections were first deparaffinized and rehydrated before antigen retrieval by heating in an appropriate buffer (citrate buffer pH 6, Life Technologies) and blocking with an IHC Blocking Solution (Enzo).

For immunofluorescence, lung tissue sections were incubated with an anti-FcεRIα primary antibody (polyclonal, Abcam), before staining with a fluorophore secondary antibody (Life Technologies). Slides were then mounted on glass slides with a DAPI-containing mounting medium, before examination on confocal microscopy (Zeiss LSM 700).

For immunohistochemistry, lung tissue sections were incubated with 3% H2O2 (Abcam) in order to block endogenous peroxidases, before incubation with an anti-FcεRIα (clone MAR-1, eBioscience), an anti-c-kit (clone 2B8, Biolegend) and an anti-Mcpt-8 (clone TUG8, Biolegend) primary antibody.

Slides were then stained with a HRP polymer-conjugated secondary antibody (ab214882, Abcam) or a biotinylated secondary antibody (ab7096, Abcam) before incubation with streptavidin-HRP (Abcam). A DAB working solution (Enzo) was used and tissue sections were counterstained with hematoxylin, dehydrated, mounted on glass slides and dried before examination with a Leica DM2000 LED microscope.

### Flow cytometry and cell sorting

Blood obtained from Townes mice was centrifuged and basophils were stained and analyzed by conventional flow cytometry in a FACSCanto II (Becton Dickinson) or Gallios (Beckman-Coulter) using FlowJo software. The following labeled mice antibodies were used: eF450 anti-CD45 (clone 30-F11, eBioscience), PerCP-eF710 anti-CD117 (clone 2B8, eBioscience), Alexa Fluor 488 anti-CD107a (clone 1D4B, eBioscience), APC anti-CD200R3 (clone Ba13, Sony), PE anti-CD49b (clone DX5, eBioscience), Pe-Cy7 anti-FcεRIα (clone MAR1, Sony). A Live/Dead^TM^ fixable Aqua viability dye (ThermoFisher Scientific) was used to exclude dead cells.

Peripheral blood mononuclear cells (PBMCs) from patients and controls were isolated by layering freshly-collected blood samples over Ficoll-Paque Plus (GE Healthcare). Conventional Flow cytometry was performed in a FACSCanto^TM^ II (Becton Dickinson) or Gallios (Beckman-Coulter) using FlowJo software. The following labeled human antibodies were used: eF506 anti-CD45 (clone HI30, eBioscience), eF450 anti-FcεRIα (clone AER-37, eBioscience), PE anti-CD123 (clone 6H6, eBioscience), APC-Cy7 anti-CCR3 (clone 5E8, BioLegend®), APC anti-CD203c (clone NP4D6, eBioscience) and FITC anti-CD63 (clone H5C6, BioLegend®).

Hyperspectral cytometry was performed in a spectral cell analyzer SP 6800 (Sony). Fluorochrome-conjugated monoclonal antibodies used for flow cytometry included: Alexa Fluor 532 anti-CD45 (clone HI30, Life Technologies), BV450 anti-FcεRI (clone AER-37, Life Technologies), PE-Dazzle594 anti-CD123 (clone 6H6, Sony), APC-Cy7 anti-CD193 (clone 5E8, Sony), BV711 anti-CD11b (clone ICRF44, Sony), PE anti-MRGPRX2 (clone K125H4, Sony), APC anti-NK1R (clone 694501, R&D), BV605 anti-CD192 (clone K036C2, Sony), PerCP-Cyanine5.5 anti-FPR1 (clone W15086B, Sony), PE-Cy7 anti-CD69 (clone FN50, Sony), BV570 anti-CD14 (clone M5E2, Sony), FITC anti-CD49a (clone TS2/7, Sony).

Human basophils were electronically sorted as CD45^+^ CD123^+^ CCR3^+^ CD203^+^ cells (in order to avoid activation by FcεRI staining), using a FACSAria II cell sorter (Becton Dickinson).

### Real-time PCR

Total RNA was extracted from sorted basophils, using the miRNeasy Micro Kit (Qiagen), and was quantified with a Nanodrop 2000 spectrophotometer (Thermo Scientific). RNA quality was assessed by 260/280 nm. Synthesis and preamplification of cDNA from basophil RNA samples were performed, using the RT2 PreAMP cDNA Synthesis kit (Qiagen). Real-time PCR was performed, using the RT² Profiler^TM^ PCR Array Human Chemokines & Receptors (Qiagen). Data were analyzed, using the ΔΔCT method.

### Murine bone-marrow-derived basophils and mast cells

Bone marrow-derived basophils were obtained by culturing bone marrow cells from femurs and tibias of C57BL/6 mice, for 1 week, in RPMI medium supplemented with fetal bovine serum (10%), nonessential amino acids (1%), penicillin/streptomycin (1%), sodium pyruvate (1 mM), 2-mercaptoethanol (54 mM; Life Technologies) and recombinant murine IL-3 (10 ng/ml; Miltenyi). Basophils were then sorted with the EasySep^TM^ Mouse CD49b Positive Selection Kit (STEMCELL Technologies), following the manufacturer’s instructions. Cell suspensions contained more than 98% basophils, as assessed by FACS analysis for CD49b^+^FcεRI^+^ expression.

Bone marrow-derived mast cells were obtained by culturing bone marrow cells from femurs and tibias of C57BL/6 mice, for 4-6 weeks, in the same medium than for basophils but with adjunction of recombinant murine SCF (10 ng/ml; Miltenyi).

### Degranulation

Murine bone marrow-derived mast cells and basophils (1×10^6^/ml) were incubated with Alexa Fluor 488 conjugate avidin (5µg/ml, Thermo Fisher Scientific) for 30 minutes, with or without substance P (10^-6^ M, Sigma), and the percentage of avidin+ cells was measured by flow cytometry.

### Chemoattraction

24-well microchemotaxis chambers (Falcon^TM^ HTS multiwell insert system, Thermo Fisher Scientific) were used and 100 µL of the basophil suspension (5×10^6^ basophils/ml) was added to the upper-compartment wells, which were separated from the lower-compartment wells by an 8 µm pore sized membrane filter (Thermo Fisher Scientific). 600 µl of LPS (10 pg/ml), histamine (10^-5^ M, Sigma) or substance P (10^-6^ M, Sigma) was added to the lower-compartment wells, allowing basophil migration during 12 hours at 37°C. A NK1R antagonist (Aprepitant, 10^-5^ M, Tocris) was used to confirm the role of this receptor in mediating substance P chemoattractant effect. The number of migrating basophils found in the lower-compartment wells was assessed by electronic count (Scepter^TM^ handheld automated cell counter, Millipore). The chemotactic index was defined as the ratio between the number of basophils found in the lower-compartment wells, after 12 hours, with the drug and without.

### Statistical analysis

Data are expressed as median [interquartile range (IQR)], or percentage. Differences between groups were assessed by Mann-Whitney test or unpaired Student’s *t* test as appropriate. Survival rates were compared by log-rank (Mantel-Cox) test. *P* < 0.05 was considered statistically significant. Data were analyzed using GraphPad Prism (GraphPad Software).

## Author contributions

S.A., R.R.-B, and T.T.M. designed the study, performed the experiments, analysed, interpreted data and wrote the manuscript.

O.H. designed the study, supervised the project and assisted in writing the manuscript. C.I., G.C. and M.B. performed and analysed some experiments.

S.A., S.M., A.S., R.F., V.B., J.B., M.M., C.H., and S.G.-L. recruited patients.

C.I., G.C., J.-B.A., V.B., J.B., M.M. J.-M.L., and S.G.-L. helped in the discussions. J.-ML. and J.C. performed histamine and substance P measurements.

## Data Availability

All data produced in the present study are available upon reasonable request to the authors

## Acknowledgements

This work was supported by state funding from the Agence Nationale de la Recherche under the Investissements d’avenir program (ANR-10-IAHU-01), the Fondation Bettencourt Schueller, the Recherche Hospitalo-Universitaire en Santé (RHU) SICKMAST and the France 2030 program through the Idex Université Paris Cité, InIdex GR-Ex (ANR-18-IDEX-0001). The authors thank the Laboratory of Excellence GR-Ex. They also thank Nicolas Montcuquet and Martin Chalumeau for helpful discussions.

