## Supplementary Table 1 for "Substance P, mast cells and basophils are involved in acute chest syndrome in sickle cell disease"

**Supplementary Table 1. Main clinico-biological characteristics of patients with SCD at steady state and during hospitalization for vaso-occlusive crisis (VOC) or acute chest syndrome (ACS)**

|  | **Steady state**  **(n = 132)** | **VOC**  **(n = 71)** | **ACS**  **(n = 30)** |
| --- | --- | --- | --- |
| Age (years) | 21.7 [14.4-33.0] | 18.6 [11.9-27.5] | 19.0 [12.3-26.3] |
| Sex (female/male) | 75/57 | 41/71 | 16/30 |
| SCD type (SS/Sβ^0^/SC) | 111/3/18 | 64/3/4 | 28/1/1 |
| Hydroxyurea | 53/132 (40.2%) | 39/71 (54.9%) | 13/30 (43.3%) |
| MET program | 35/132 (26.5%) | 1/71 (1.4%) | 1/30 (3.3%) |
| Hemoglobin level (g/dL) | 9.4 [8.4-10.5] | 8.4 [7.7-9.3] | 7.8 [7.2-9.1] |
| Reticulocyte count (G/L) | 243 [163-348] | 263 [192-353] | 312 [139-377] |
| Leukocyte count (G/L) | 8.8 [6.7-12.4] | 11.5 [9.4-13.6] | 10.5 [7.4-17.4] |
| Neutrophil count (G/L) | 4.6 [3.1-6.6] | 6.4 [4.5-8.9] | 5.9 [4.1-12.3] |
| Platelet count (G/L) | 355 [274-454] | 402 [261-483] | 307 [190-417] |
| LDH level (U/L) | 375 [318-457] | 472 [290-544] | 475 [369-618] |
| AST level (U/L) | 43 [33-52] | 49 [40-66] | 41 [35-57] |
| Free bilirubin level (µmol/L) | 30 [17-46] | 27 [19-34] | 27 [19-46] |
| CRP level (mg/L) | 3.4 [1.9-5.7] | 45 [16-101] | 85 [65-185] |

Data are median [interquartile range] or percentage.

ACS: acute chest syndrome. AST: aspartate aminotransferase. CRP: C-reactive protein. LDH: lactate dehydrogenase. MET: monthly exchange transfusion. SCD: sickle cell disease. VOC: vaso-occlusive crisis.
